# Application of SinoPlan in Trajectory Planning for Robot-Assisted Intracerebral Hematoma Puncture: A Single-Centre Retrospective Comparative Study

**DOI:** 10.64898/2026.05.24.26353998

**Authors:** Hu An, Gu Yu, Zhou Qiangyi, Fang Chaoyou, Wu Xiang, Wang Yan, Ding Wei, Yao Jian, Zhang Yunfeng

## Abstract

Robot-assisted hematoma puncture has seen significant development in primary hospitals across the country. Sino Plan software system is the core of the intelligent surgical robot, independently developed by Sinovation.We conducted a comparative study of imaging indicators, such as residual hematoma volume and hematoma clearance rate, as well as prognostic indicators, in patients who underwent hematoma puncture at our hospital over a 9-year period, before and after the introduction of Sino Plan.The results indicated that following the application of Sino Plan, the hematoma clearance rate was significantly enhanced, and the residual hematoma volume was markedly reduced. Regarding patient prognosis, there was no significant difference in GCS scores between the two groups, but the incidence of adverse prognostic events was lower in patients where Sino Plan was utilized.In conclusion, this 9-year retrospective analysis at our hospital reveals that Sino Plan offers distinct advantages. However, its application in certain special cases suggests that further improvements to the software are warranted to better meet the demands of more specific clinical scenarios.

**Background:** Intracranial hematoma remains a major cause of death and disability worldwide. Minimally invasive stereotactic or robot-assisted hematoma puncture has become increasingly used as an alternative to conventional craniotomy, but its effectiveness depends critically on accurate trajectory planning. SinoPlan is a neurosurgical planning platform that supports multimodal image fusion, lesion segmentation, three-dimensional reconstruction, and trajectory optimisation. We aimed to evaluate the clinical value of SinoPlan in hematoma puncture for ICH in real-world practice.

**Methods:** In this single-centre retrospective comparative study, we reviewed patients who underwent intracerebral hematoma puncture at Anqing 116 Hospital between February, 2017, and February, 2025. Patients treated before the introduction of SinoPlan were assigned to the control group, and those treated after its implementation in February, 2023, were assigned to the SinoPlan group. Radiological outcomes, including hematoma clearance rate and residual hematoma volume, were compared between groups. Subgroup analyses were performed according to hematoma volume and morphology. Clinical outcomes, including Glasgow Coma Scale (GCS) scores, modified Rankin Scale (mRS), and adverse events, were also assessed.

**Results:** A total of 80 patients were included, with 55 in the control group and 25 in the SinoPlan group. Compared with conventional planning, SinoPlan-assisted trajectory planning was associated with a significantly higher hematoma clearance rate, a significantly lower residual hematoma volume, and better trajectory optimisation. These benefits were observed in both hematomas smaller than 30 mL and those larger than 30 mL. In subgroup analyses by morphology, SinoPlan improved trajectory optimisation in both regular and irregular hematomas, although the radiological benefit appeared less pronounced in irregular lesions. No significant between-group differences were found in serial GCS scores or mRS distributions. However, adverse prognostic events were less frequent in the SinoPlan group, and no deaths occurred within 3 months in patients treated with SinoPlan-assisted surgery.

**Conclusion:** SinoPlan-assisted trajectory planning improved radiological evacuation outcomes and was associated with fewer adverse events in intracerebral hematoma puncture. These findings support the clinical utility of software-assisted planning in minimally invasive ICH surgery, while also suggesting that further optimisation may be needed for anatomically irregular hematomas.

## Introduction

Intracranial hematoma is a major cause of death and disability worldwide^[1, 2]^. Intracranial hematoma usually includes spontaneous intracerebral hemorrhage and traumatic hematoma.Although advances in critical care have improved perioperative management, outcomes remain poor, especially in patients with deep hematomas, mass effect, and secondary neurological deterioration^[3]^. Surgical evacuation is biologically plausible because removal of clot burden may relieve local compression and reduce secondary injury related to ischemia, inflammation, and blood-product toxicity^[4, 5]^. Yet the role of surgery in Intracranial hematoma remains unsettled, and traditional craniotomy has not consistently shown functional benefit in large clinical trials.

Minimally invasive hematoma evacuation has emerged as an alternative strategy aimed at reducing surgical trauma while achieving effective clot reduction. Among these techniques^[6]^, stereotactic puncture and catheter drainage are increasingly used because they are relatively simple, rapid, and reproducible. More recently, the incorporation of robotic and stereotactic technologies has improved procedural precision and expanded the feasibility of minimally invasive evacuation in routine practice^[7-9]^. However, clinical success depends not only on the procedure itself but also on accurate trajectory planning. An optimal trajectory should reach the core or long axis of the hematoma while avoiding vessels, eloquent cortex, ventricular structures, and major white-matter pathways^[10-12]^. This requirement is particularly important in deep or small-volume hematomas, in which small targeting errors may substantially compromise efficacy or safety.

Sinoplan™ software planning system, independently developed by Sinovation®, is the soul of the intelligent surgical robot. With continuous innovation over the past decade, Sinoplan™ surgical planning system has now obtained Class III medical device registration approval.The SinoPlan surgical planning software can fuse, segment, and reconstruct different types of patient medical images, presenting them comprehensively and intuitively before you, assisting you in easily developing safe and precise surgical plans. As the soul of the surgical robot, SinoPlan incorporates more advanced artificial intelligence imaging algorithms and has always been closely aligned with the clinical needs and usage habits of Chinese doctors, continuously evolving.

We therefore conducted a retrospective comparative study of patients who underwent intracranial hematoma puncture at our centre between 2017 and 2025, before and after the introduction of SinoPlan into clinical workflow. We aimed to determine whether SinoPlan-assisted planning was associated with improved hematoma evacuation, reduced residual hematoma volume, and better clinical safety, and to examine whether these effects varied according to hematoma volume and morphology.

## Methods

### Study design and participants

This was a single-centre retrospective comparative study conducted at An qing 116 Hospital. We reviewed consecutive patients who underwent minimally invasive intracerebral hematoma puncture and drainage between Feb 1, 2017, and Feb 29, 2025. Patients were divided into two groups according to the surgical planning strategy used during the study period. The control group comprised patients who underwent surgery between Feb 1, 2017, and Feb 1, 2023, before the routine introduction of SinoPlan into clinical workflow. The SinoPlan group comprised patients who underwent surgery between Feb 1, 2023, and Feb 29, 2025, after implementation of SinoPlan-assisted preoperative planning.

Eligible patients met all of the following criteria: (1) intracerebral hematoma confirmed by CT or MRI; (2) symptoms or signs suggestive of increased intracranial pressure, including headache, nausea, vomiting, or impaired consciousness; (3) evidence of mass effect with midline shift and a general condition considered suitable for surgery; (4) no severe bleeding tendency, active infection, or other absolute contraindication to surgery; and (5) provision of informed consent by the patient or their legal representative.

Patients were excluded if they had: (1) hematoma secondary to vascular malformation, ruptured aneurysm, or other structural lesions requiring alternative treatment; (2) established cerebral herniation considered unlikely to benefit from puncture and drainage alone; (3) severe coagulopathy, including marked thrombocytopenia or coagulation factor deficiency; (4) systemic or intracranial infection; (5) poor general condition precluding surgery; (6) brain death or end-stage neurological injury; (7) hematoma located in a critical functional region or with an excessively high-risk trajectory judged unsuitable for puncture; or (8) refusal of surgery by the patient or family.

This study was approved by the institutional review board of our hospital. Given the retrospective nature of the study, the requirement for additional written informed consent for study participation was waived where permitted by local regulations.

### Data collection

Clinical and imaging data were retrieved from the hospital electronic medical record system and radiological archive. Baseline variables included age, sex, date of admission, side and location of hematoma, preoperative hematoma volume, hematoma morphology, preoperative Glasgow Coma Scale (GCS) score, and relevant perioperative information.

Operative records were reviewed to identify the planning modality, puncture approach, entry point, trajectory direction, use of stereotactic assistance, and perioperative complications. Follow-up clinical data included GCS score on postoperative day 1, postoperative day 30, and postoperative day 90, as well as modified Rankin Scale (mRS) score at 1 month and 3 months after surgery when available. Mortality and adverse prognostic events within 3 months were also recorded.

Radiological data were collected from preoperative and postoperative CT scans. Preoperative imaging was used to determine hematoma location, volume, shape, and planned puncture trajectory. Postoperative imaging was used to assess residual hematoma volume and hematoma clearance rate.

### Surgical planning and procedure

All patients underwent minimally invasive hematoma puncture and drainage using a stereotactic localisation device according to routine institutional practice. In the control group, surgical planning was based on conventional review of preoperative imaging by the operating surgeon. Entry point and trajectory were selected using standard axial and multiplanar CT assessment, with the aim of reaching the central or dependent portion of the hematoma while avoiding obvious cortical vessels, ventricular penetration when possible, and eloquent brain regions.

In the SinoPlan group, preoperative planning was performed using the SinoPlan software platform. Multimodal imaging data, including CT and MRI when available, were imported into the system for image fusion and registration. The hematoma was delineated using the software-assisted lesion contouring tools, and a three-dimensional model of the lesion was reconstructed. Where available, vascular structures, brain surface anatomy, and relevant fibre pathways or functional regions were also reconstructed or visualised. Candidate puncture trajectories were then simulated, and the final path was selected on the basis of maximising access to the long axis or core of the hematoma while minimising transgression of vessels, functional cortex, and critical white matter structures.

After preoperative planning, the stereotactic device was mounted, and intraoperative data acquisition was completed according to the standard procedural workflow. The puncture site was identified on the basis of the planned entry point, and the direction of the puncture was confirmed using the stereotactic system. A drainage catheter was then advanced along the planned trajectory into the hematoma cavity. Postoperative management, including drainage and further medical treatment, followed institutional protocols and was similar in both groups.

### Image analysis and trajectory assessment

Preoperative hematoma volume and postoperative residual hematoma volume were measured from CT imaging. When available, lesion segmentation in the SinoPlan group was performed using the software contouring tools; in the control group, volume assessment was based on routine radiological measurement. Hematoma clearance rate was calculated as:

**hematoma clearance rate = (preoperative hematoma volume – postoperative residual hematoma volume) / preoperative hematoma volume × 100%**.

Hematomas were further classified according to volume and morphology for subgroup analysis. Based on preoperative volume, patients were stratified into hematomas smaller than 30 mL and hematomas 30 mL or larger. Based on imaging appearance, hematomas were classified as regular-shaped or irregular-shaped according to the contour of the lesion on preoperative imaging.

Trajectory quality was assessed by comparing the actual or planned puncture path with the presumed optimal trajectory to the hematoma. In our analysis, the ratio between the distance from the puncture endpoint to the long axis of the hematoma and the short axis of the hematoma was used as an index of trajectory optimisation, with a higher value indicating closer approximation to the optimal path. In addition, representative cases were reviewed qualitatively to illustrate how SinoPlan altered conventional path selection in selected deep or anatomically complex hematomas.

### Outcomes

The primary outcomes were radiological efficacy measures: postoperative residual hematoma volume and hematoma clearance rate.

Secondary outcomes included trajectory optimisation, proportion of patients achieving effective hematoma evacuation, proportion achieving satisfactory hematoma evacuation, and subgroup effects according to hematoma volume and morphology. Clinical secondary outcomes included GCS score before surgery and at postoperative day 1, day 30, and day 90, as well as mRS score at 1 month and 3 months. Safety outcomes included adverse prognostic events within 3 months after surgery and all-cause mortality during follow-up.

Where applicable, effective and satisfactory hematoma evacuation were defined according to prespecified institutional thresholds based on postoperative radiological reduction in hematoma burden. Adverse prognostic events included death and other clinically significant postoperative deterioration events recorded during follow-up.

### Statistical analysis

All statistical analyses were performed using GraphPad software. Continuous variables were first assessed for distributional characteristics. If data followed a normal distribution, between-group comparisons were performed using Student’s t test. If data did not follow a normal distribution, the rank-sum test was used. Categorical variables were compared using the χ^2^ test or Fisher’s exact test, as appropriate.

Data are presented as mean (SD), median (IQR), or number (%), as appropriate. Subgroup analyses were performed according to hematoma volume (<30 mL vs ≥30 mL) and hematoma morphology (regular vs irregular). All tests were two-sided, and a p value of less than 0·05 was considered statistically significant.

## Results

### Patient characteristics

A total of 80 patients were included in the final analysis, including 55 patients in the control group and 25 patients in the SinoPlan group. Patients in the control group underwent surgery before the routine implementation of SinoPlan-assisted planning, whereas those in the SinoPlan group were treated after its introduction into clinical practice. Baseline demographic, radiological, and perioperative data were collected retrospectively from medical records and imaging archives for comparative analysis. Please refer to the table below for specific patient information (Table 1).

**Table 1:** Clinical Information of 90 Patients Undergoing Hematoma Evacuation at Anqing 116 Hospital.

| Characteristic | Overall | Ctrl | Sino | p-value |
| --- | --- | --- | --- | --- |
| Case (n) | 90 | 60 | 30 | / |
| Age (year) | 59.83 ± 16.73 | 59.05 ± 17.91 | 61.4 ± 14.24 | 0.5329 |
| Sex (male/female) | 57/33 | 36/24 | 21/9 | / |
| Operation time | 2017.1.1-2025.12.31 | 2017.1.1-2023.2.1 | 2023.2.1-2025.12.31 | / |
| Supratentorial Hematoma (n) | 80 | 55 | 25 | / |
| Hematoma volume(> 30ml) (n/percentage(%)) | 49/61.3% | 35/63.6% | 14/56% | / |
| Regular shape(n/percentage(%)) | 57/71.3% | 42/76.4% | 15/60% | / |

### SinoPlan enabled three-dimensional preoperative planning and optimised surgical trajectory selection

SinoPlan provided an integrated planning platform for intracerebral hematoma puncture, incorporating lesion contouring, multimodal image fusion, three-dimensional reconstruction, vascular visualisation, and fibre bundle reconstruction (Figure 1A). These functions were embedded into the standard stereotactic puncture workflow, including device mounting, data acquisition, puncture site localisation, and intraoperative confirmation of trajectory direction (Figure 1B).

**Figure 1:**
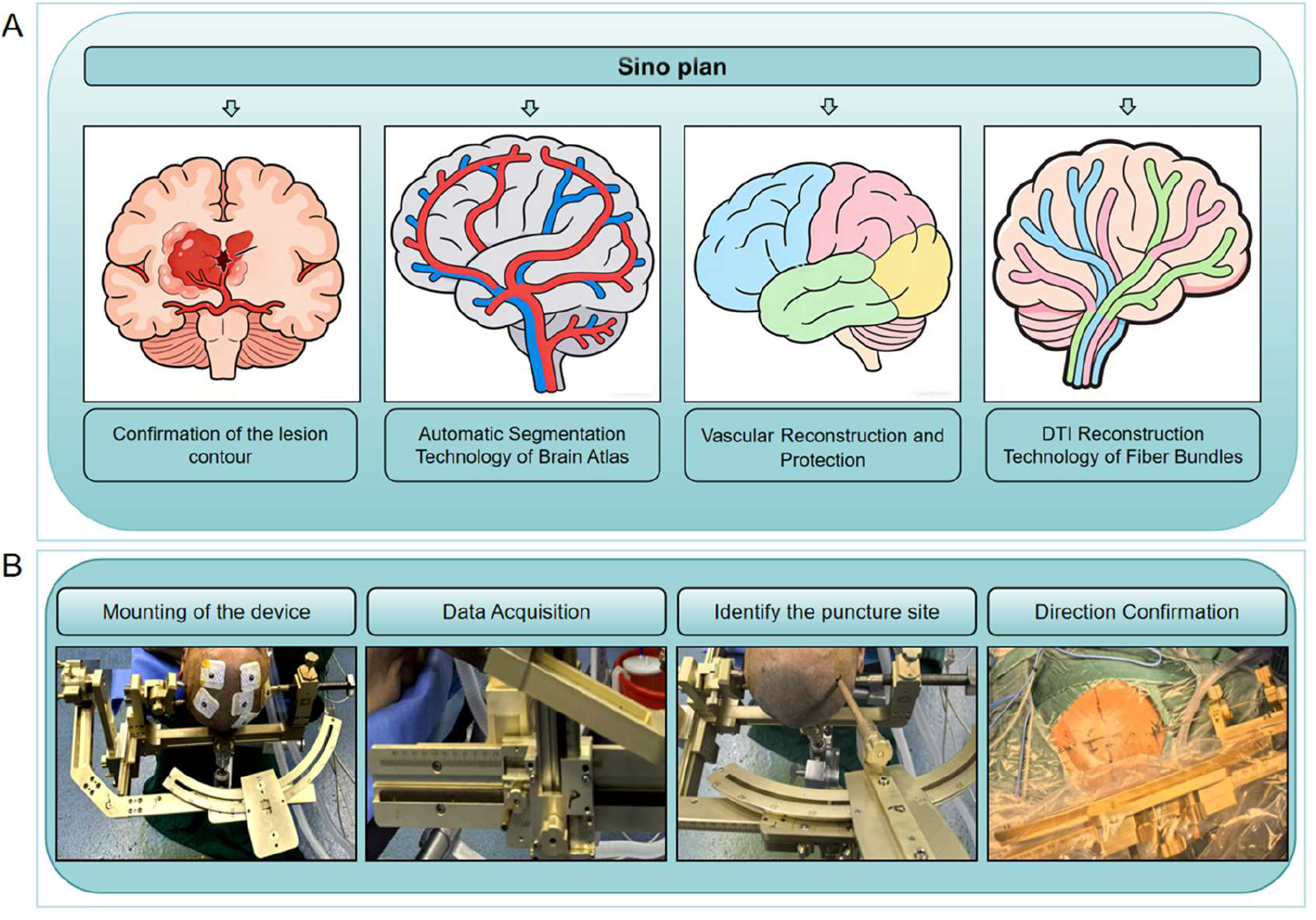
SinoPlan-assisted workflow for minimally invasive puncture and drainage of intracerebral haematoma. SinoPlan-assisted preoperative planning and operative workflow. **(A)** Core functions of the SinoPlan platform, including lesion contouring, multimodal image fusion, three-dimensional reconstruction, vascular visualisation, and diffusion tensor imaging-based fibre tracking. **(B)** Standard workflow for stereotactic minimally invasive puncture and drainage, including device mounting, data acquisition, puncture site localisation, and confirmation of puncture direction.

Representative cases further illustrated the practical role of SinoPlan in preoperative planning (Figure 2). Through three-dimensional anatomical simulation, SinoPlan enabled more precise localisation of deep or small hematomas and facilitated optimisation of the entry point and trajectory. In selected cases, SinoPlan-assisted planning altered the original surgical approach by providing a more direct and anatomically safer path to the hematoma cavity. Postoperative imaging in these representative patients confirmed satisfactory catheter placement and hematoma evacuation. Together, these findings suggest that SinoPlan can improve the visualisation and planning of minimally invasive hematoma puncture in routine clinical practice.

**Figure 2:**
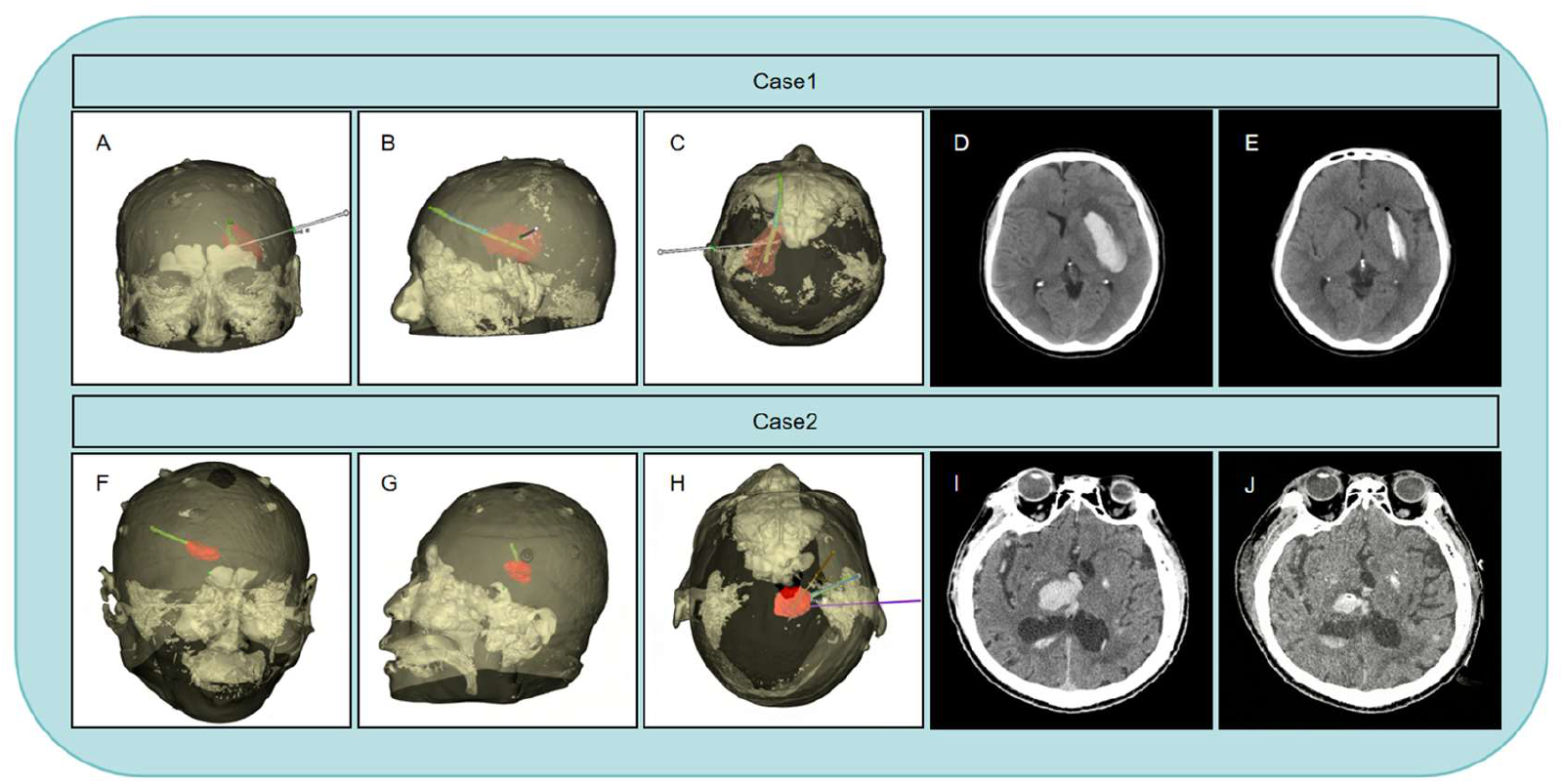
Representative cases of SinoPlan-assisted trajectory planning. Representative cases showing three-dimensional reconstruction and trajectory planning with SinoPlan for deep intracerebral haematoma puncture. **(A–E)** Case 1. Three-dimensional views of the planned trajectory and corresponding preoperative and postoperative CT images. **(F–J)** Case 2. Three-dimensional views of the planned trajectory and corresponding preoperative and postoperative CT images. SinoPlan enabled preoperative simulation of the puncture route and optimisation of the surgical approach.

### SinoPlan improved radiological evacuation outcomes across different hematoma subgroups

We performed a statistical analysis of the imaging data of supratentorial hematoma. Compared with conventional planning, SinoPlan-assisted procedures achieved significantly better radiological outcomes (Figure 3). Overall, the SinoPlan group showed a significantly higher trajectory optimisation index, a significantly greater hematoma clearance rate, and a significantly lower residual hematoma volume than the control group (Figure 3A–C). In addition, the proportions of patients achieving effective hematoma evacuation and satisfactory hematoma evacuation were higher in the SinoPlan group (Figure 3D, E).

**Figure 3:**
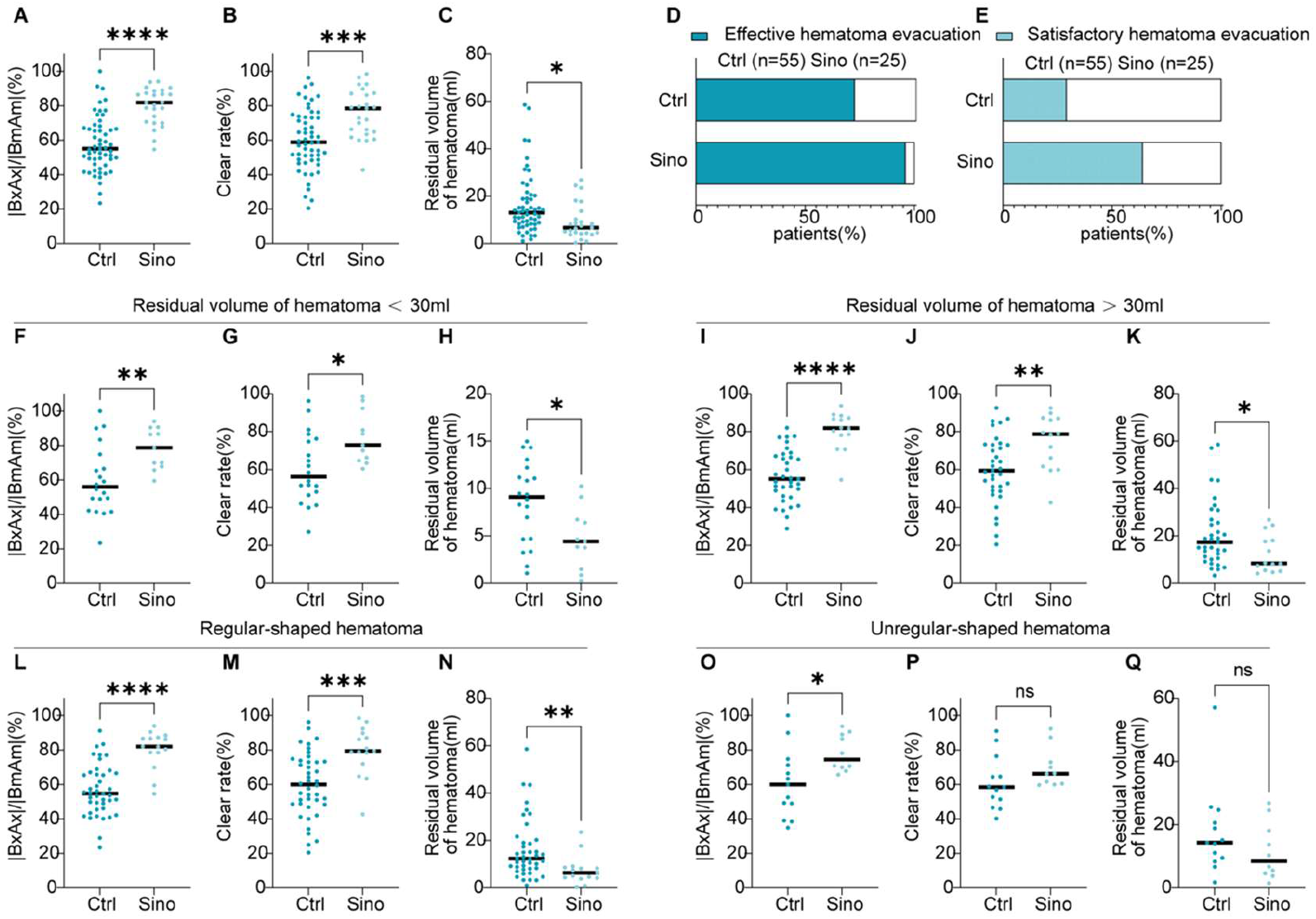
SinoPlan-assisted planning improved trajectory optimisation and radiological evacuation outcomes. Comparison of radiological outcomes between the control group and the SinoPlan group. **(A–E)** Overall comparisons of trajectory optimisation index, haematoma clearance rate, residual haematoma volume, and proportions of effective and satisfactory evacuation. **(F–K)** Subgroup analyses according to haematoma volume. **(L–Q)** Subgroup analyses according to haematoma morphology. Data are mean (SD), median (IQR), or n (%), as appropriate. *p* values were calculated using Student’s *t* test, rank-sum test, as appropriate. ns=not significant.

Subgroup analyses according to hematoma volume showed that these advantages were maintained in both smaller and larger hematomas. In patients with hematoma volume less than 30 mL, SinoPlan was associated with improved trajectory optimisation, increased hematoma clearance, and reduced residual hematoma volume (Figure 3F–H). Similar findings were observed in patients with hematoma volume greater than 30 mL (Figure 3I–K), indicating that the radiological benefit of SinoPlan was not restricted to a specific lesion size.

When patients were stratified by hematoma morphology, SinoPlan also demonstrated superior trajectory optimisation in both regular and irregular hematomas. In regular-shaped hematomas, SinoPlan significantly improved the trajectory optimisation index, hematoma clearance rate, and residual hematoma volume compared with conventional planning (Figure 3L–N). In irregular-shaped hematomas, SinoPlan still significantly improved the trajectory optimisation index, although no significant between-group differences were found in hematoma clearance rate or residual hematoma volume (Figure 3O–Q). These findings suggest that SinoPlan consistently improved geometric trajectory planning, but its radiological advantage was more pronounced in regular than in irregular hematomas.

### SinoPlan was associated with fewer adverse events, although short-term neurological outcomes were comparable between groups

Clinical outcome analysis showed no significant between-group differences in Glasgow Coma Scale (GCS) scores before surgery, on postoperative day 1, postoperative day 30, or postoperative day 90 (Figure 4A–D). Likewise, the distribution of GCS-based recovery categories at 3 months was not significantly different between groups (Figure 4E). Functional outcomes assessed by modified Rankin Scale (mRS) at 1 month and 3 months were also broadly similar between groups (Figure 4F, G).

**Figure 4:**
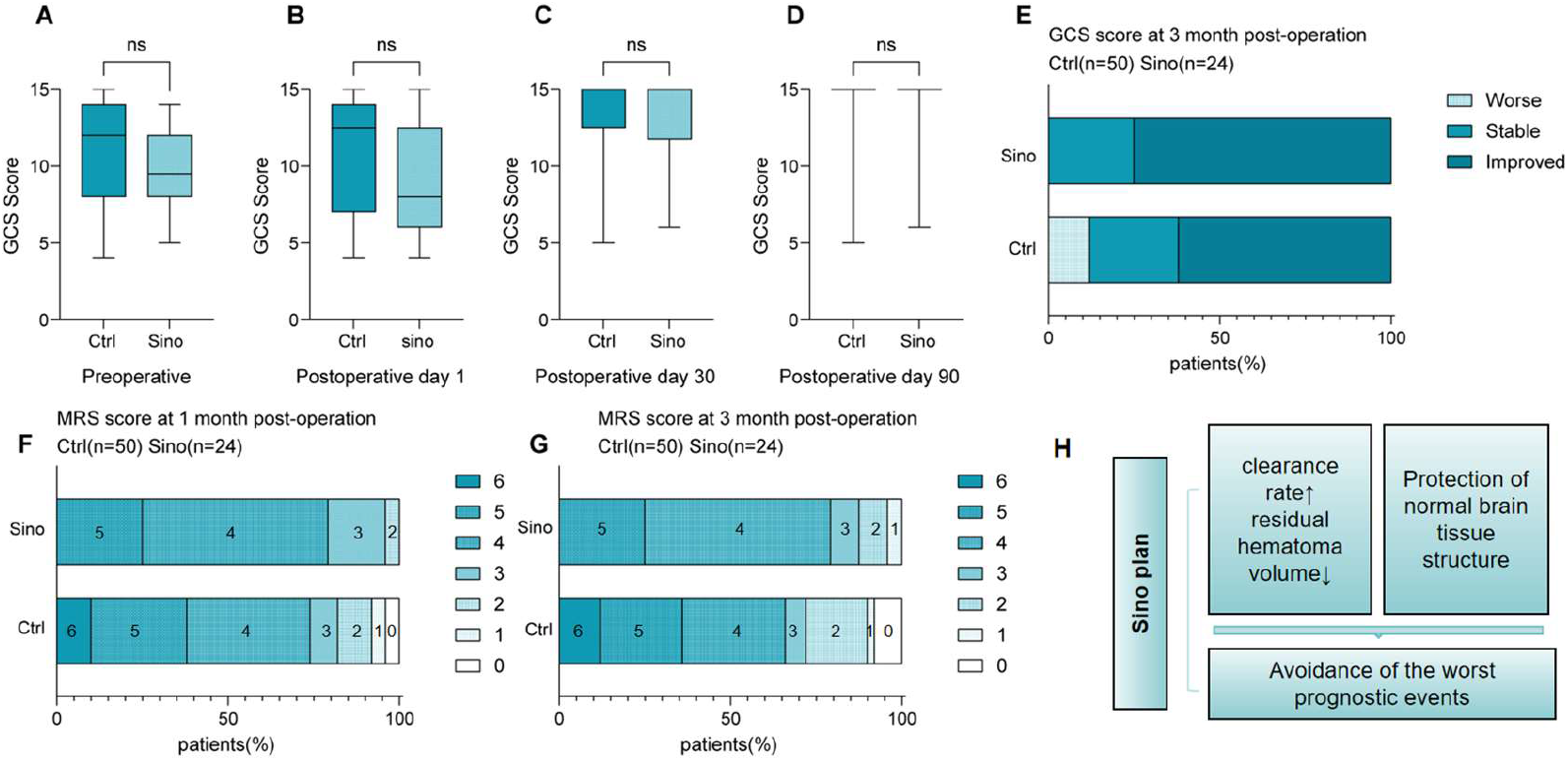
Clinical outcomes after conventional versus SinoPlan-assisted intracerebral haematoma puncture. Comparison of clinical outcomes between the control group and the SinoPlan group. **(A–D)** Glasgow Coma Scale scores before surgery and at postoperative day 1, day 30, and day 90. **(E)** Distribution of 3-month Glasgow Coma Scale-based outcome categories. **(F, G)** Modified Rankin Scale scores at 1 month and 3 months after surgery. Data are mean (SD), median (IQR), or n(%), as appropriate. *p* values were calculated using Student’s *t* test, rank-sum test, as appropriate. ns=not significant.**(H)** Illustration of the application conclusions of the Sino Plan

However, despite the lack of significant differences in short-term neurological scale scores, adverse prognostic events were less frequent in the SinoPlan group. Notably, no deaths occurred within 3 months among patients who underwent SinoPlan-assisted surgery. These findings indicate that the main benefit of SinoPlan in this cohort was reflected in improved radiological evacuation and perioperative safety rather than in measurable short-term differences in neurological recovery scales.

## Discussion

In this single-centre retrospective study, SinoPlan-assisted preoperative planning improved the radiological performance of minimally invasive intracerebral haematoma puncture compared with conventional planning. Specifically, the SinoPlan group showed better trajectory optimisation, higher haematoma clearance rates, lower residual haematoma volumes, and higher proportions of effective and satisfactory evacuation. These findings support the value of software-assisted planning in improving the technical accuracy and evacuation efficiency of minimally invasive surgery for intracerebral haemorrhage.

These results are consistent with the growing body of literature supporting minimally invasive evacuation as an important treatment strategy for intracerebral haemorrhage, particularly when conventional craniotomy may cause substantial surgical trauma. Previous studies have shown that the effectiveness of minimally invasive puncture depends heavily on accurate selection of the entry point and trajectory^[13]^, as suboptimal catheter placement can result in insufficient drainage, greater postoperative residual haematoma, repeated adjustment, and unintended injury to surrounding brain tissue^[14-16]^. In this context, image-guided and software-assisted planning systems have attracted increasing attention as tools to improve the precision and reproducibility of haematoma evacuation^[17]^. Our findings extend this concept by showing, in a real-world clinical setting, that a practical neurosurgical planning platform can improve not only trajectory-related indices but also final radiological evacuation outcomes.

The observed benefit of SinoPlan is likely related to its ability to provide intuitive three-dimensional visualisation of the haematoma and surrounding anatomy, allowing surgeons to identify a more appropriate entry point and puncture trajectory. By integrating lesion reconstruction with anatomical and vascular information, the platform may help define a route that more directly accesses the target while avoiding important adjacent structures. This advantage may be especially relevant in deep-seated haematomas, in which even small deviations in trajectory can substantially affect catheter placement and drainage efficacy. The representative cases in our study also suggest that SinoPlan may influence surgical decision making by enabling preoperative simulation and optimisation of the operative approach.

Subgroup analyses further showed that the radiological benefit of SinoPlan was present in both smaller and larger haematomas, suggesting that its value was not restricted to a specific lesion size. However, when patients were stratified by haematoma morphology, the benefit appeared to differ. In regular-shaped haematomas, SinoPlan improved both trajectory-related and evacuation-related outcomes, whereas in irregular-shaped haematomas, although trajectory optimisation still improved, the advantages in haematoma clearance and residual volume were less evident. A possible explanation is that irregular haematomas often have more complex and multilobulated clot architecture, so even a well-planned single trajectory may not adequately access all compartments of the lesion. As a result, improved geometric planning may not necessarily translate into proportionally better drainage in these cases.

Despite the radiological improvements, no significant between-group differences were observed in short-term neurological or functional outcomes, including GCS and mRS scores. This is broadly in line with previous studies of intracerebral haemorrhage, in which functional recovery is determined not only by the technical success of haematoma evacuation but also by multiple other factors, including initial haematoma burden, haematoma location, secondary brain injury, age, comorbidities, perihaematomal oedema, systemic complications, and postoperative rehabilitation^[3]^. Therefore, better radiological evacuation does not always translate into measurable short-term functional superiority, particularly in studies with limited sample size and relatively short follow-up. Nevertheless, adverse prognostic events were less frequent in the SinoPlan group, and no deaths occurred within 3 months in this group, suggesting a possible safety benefit that merits further investigation.

Several limitations should be acknowledged. First, this was a retrospective single-centre study with a relatively small sample size, which may limit generalisability and introduce selection bias. Second, the two groups were defined according to different treatment periods before and after implementation of SinoPlan, so temporal changes in surgical experience or perioperative care may have influenced the results. Third, although radiological outcomes were clearly improved, the study may have been underpowered to detect differences in functional recovery. Finally, the present analysis did not determine which specific components of the planning platform contributed most to the observed benefit.

Overall, our findings suggest that SinoPlan-assisted planning can improve trajectory design and radiological evacuation quality in minimally invasive intracerebral haematoma puncture, with a possible benefit for procedural safety. Larger prospective studies are needed to confirm these findings and to determine whether improved planning can ultimately translate into better long-term neurological outcomes.

## Conclusion

SinoPlan-assisted preoperative planning improved trajectory optimisation and radiological evacuation outcomes in minimally invasive intracerebral hematoma puncture. Compared with conventional planning, SinoPlan was associated with higher hematoma clearance, lower residual hematoma volume, and fewer adverse prognostic events. The benefit was observed across hematoma-volume subgroups and appeared more pronounced in regular than in irregular hematomas. Although short-term neurological and functional outcomes were not significantly different between groups, it has the effect of reducing the incidence of extremely poor prognostic events. these findings support the value of software-assisted planning in ICH surgery and warrant further prospective validation.

## Data Availability

The data that support the findings of this study are available from the corresponding author upon reasonable request

## Abbreviations

ICH: intracerebral haemorrhage
CT: computed tomography
DTI: diffusion tensor imaging
GCS: Glasgow Coma Scale
mRS: modified Rankin Scale
SD: standard deviation
IQR: interquartile range

## Data availability statement

The datasets generated and/or analysed during the current study are not publicly available because of patient privacy and institutional restrictions, but are available from the corresponding author upon reasonable request and with permission from the relevant institution.

## Ethics statement

This retrospective study was approved by the Institutional Review Board/Ethics Committee of Anqing 116 Hospital. Written informed consent was obtained from all patients or their legal representatives.

## Author contributions

Conceptualization, [A], [B]; Methodology, [A], [B], [C]; Data curation, [D], [E]; Formal analysis, [F], [G]; Investigation, [A], [C], [D]; Visualization, [B], [E]; Writing – original draft, [A], [B],[H]; Writing – review & editing, all authors; Supervision, [I]. All authors approved the final manuscript.

